# Telemedicine for surgical site infection diagnosis: a systematic review protocol

**DOI:** 10.1101/2022.01.31.22270148

**Authors:** Ross Lathan, Misha Sidapra, Marina Yiasemidou, Judith Long, Joshua Totty, George Smith, Ian Chetter

## Abstract

1

Since the COVID-19 pandemic there has been a rapid uptake and utilisation of telemedicine in all aspects of healthcare. This presents a key opportunity in surgical site infection surveillance. Remote follow up methods have been used via telephone, with photographs and questionnaires for post-operative reviews with varying results.

This review therefore aims to comprehensively synthesise available evidence for the diagnostic accuracy of all forms of SSI telemedicine monitoring. The protocol has been established as per both PRISMA-P and the Cochrane handbook for reviews of diagnostic test accuracy.

Medline, Embase, CENTRAL and CINAHL will be searched using a complete search strategy developed with librarian input, in addition to google scholar and hand searching. All study designs with patients over 18 and undergone a primarily closed surgical procedure will be eligible. Index tests will include all forms of telemedicine and a subgroup analysis performed for each of these. Comparative tests must include face to face review, and all reference standards will be included again for sub-group analyses. Search results will be screened by two investigators independently with a third providing consensus review on disagreements. Methodological quality will be assessed using the QUADAS-2 tool, first validated by two investigators as per the Cochrane handbook.

Exploratory analysis will formulate summary receiver operating characteristic curves and forest plots with estimates of sensitivity and specificity of the included studies. Sources of heterogeneity will be identifying and investigated through further analysis.

Potential benefits of telemedicine integration in surgical practice will reduce cost and travel time to patients in addition to avoiding wasted clinic appointments, important considerations in a peri-pandemic era. To avoid missed or further complications, there must be confidence in the ability to diagnose infection. This review will systematically determine whether telemedicine is accurate for surgical site infection diagnosis, which methods are well established and if further research is indicated.

**Amendments:** Protocol version number will be annotated on the title page and thereby footer of each page. A list of major amendment changes to the protocol will be kept in the table below.

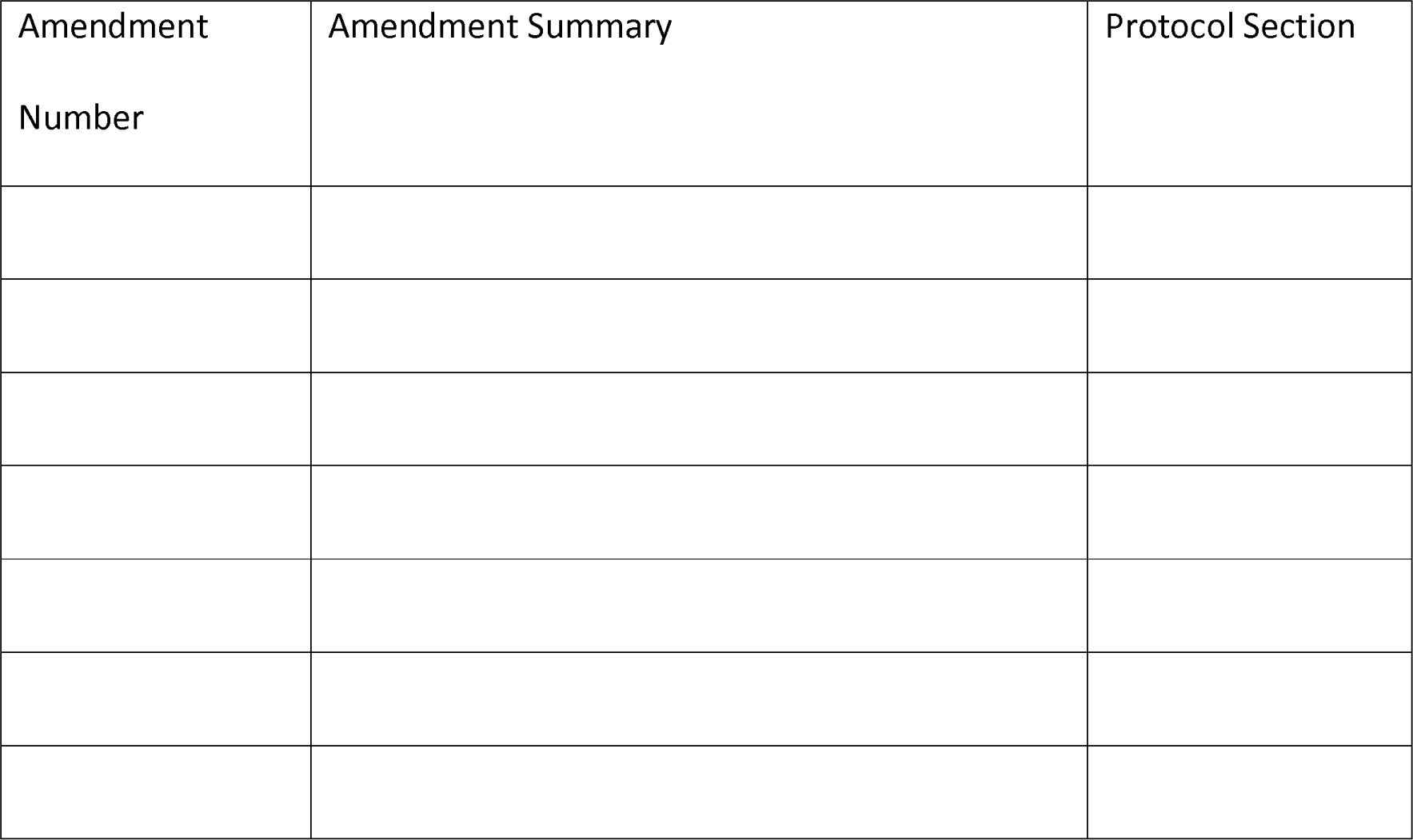

**Important Dates:** 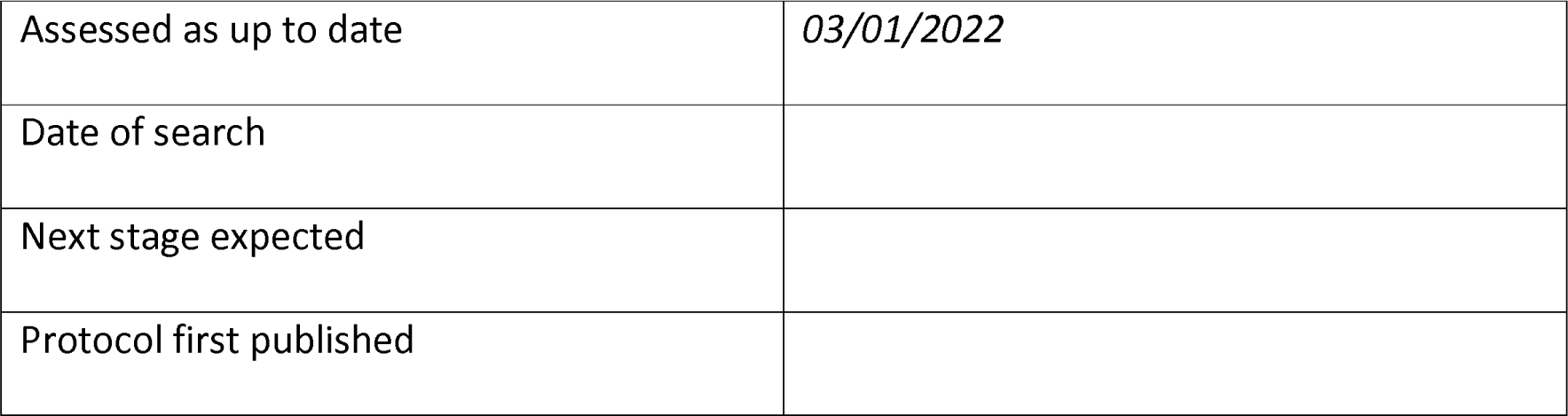

**Definitions:** Surgical Site Infection An infection occurring after surgery in the part of the body where surgery took place

Telemedicine The remote diagnosis and treatment of patients by means of communications technology

Telehealth The delivery of healthcare services, where patients and providers are separated by distance.

eHealth The use of information communication technology for health.

mHealth A component of eHealth where medical and public health practices are supported by mobile devices.

Virtual Care All the ways healthcare providers remotely interact with their patients

## 2 Background

### 2.1 Target condition being diagnosed

This review primarily aims to identify surgical site infections (SSI). The Centre for Disease Control and Prevention (CDC) defines SSI as an infection within 30 days of an operation or up to 90 days if an implant is left in place and the infection is related to an operative procedure(1). SSI are further classified as ‘superficial incisional SSI’, ‘deep incisional SSI’, and ‘organ/space SSI’; further details of these definitions can be found in appendix one(2).

Surgical site infections complicate over 30% of operations, depending on the type of procedure(3, 4). Up to 60% of these present after discharge and so accurate and timely diagnosis requires intensive follow-up and potentially significant travel distances on the patient’s behalf(5). For patients, SSI may have a significant impact on morbidity and mortality with subsequent time and cost implications(6). Unsurprisingly, the burden of infection encompasses healthcare providers too, with a recent UK study showing an association of SSI with a 92% increase in length of stay and an adjusted episode cost of £3040(7). Treatment for SSI can range from an oral course of antibiotics, to the need for reintervention (drainage or debridement), prolonged inpatient readmission and the subsequent risk of further morbidity (i.e. thrombotic and ischaemic events).

### 2.2 Index and Gold Standard Tests

The current ‘gold standard’ for diagnosing SSI is a ‘face-to-face’ review using the US CDC criteria(2). However, other scoring systems or criteria may be used during a face-to-face review. The ASEPSIS score uses weighted, objectively measurable criteria to identify surgical wounds as satisfactory healing, impaired wound healing or infected (appendix three)(8). A newly developed utility, the Bluebelle wound healing questionnaire (WHQ) has been validated in general surgery as a patient or clinician reported outcome measure to identify surgical site infection(9).

Diagnosis of SSI using remote, digitally based contact between patients and clinicians (telemedicine) is being investigated as the primary index test in this review. In this review we are focussing on digital remote follow-up. There are many ways of implementing this in practice. These include the use of photographic images and/or video, either in real time or deferred, telephone review and instant messaging. There may be other novel methods not listed here that are identified during the review process.

### 2.3 Clinical Pathway

Patients undergoing surgery may present with SSI at one of three typical time points. If an infection does occur, the first potential route of diagnosis is prior to discharge and within the first week. Infections at this point will depend on the surgical procedure, underlying comorbidities, and age, as these factors will influence typical length of stay (LOS) and wound healing. They are likely to be picked up on ward rounds or through dressing changes with the nursing team then highlighting issues to the surgeon. Face to face diagnosis using CDC criteria in this instance is straightforward as the patient has not left the department. The patient may have to undergo further observation, dressing changes and antibiotic course. Rarely, further imaging or intervention may be required.

The second point is typically patient initiated, after discharge but prior to any planned follow up, within 30 days of operation. Infections at this stage may not reach the surgeon’s knowledge as they are often managed by the patient’s primary care physician. However, without specialist input some infections progress in severity. Patients with evidence of deep incisional or organ/space infections may be referred to secondary care by the primary care team, may present to the emergency department or may contact the surgical team directly through an aftercare number. Only once seen can the gold standard assessment take place. Further imaging, microbiology culture and sensitivity testing are often implemented to ensure appropriate and specific management. Further surgical intervention may be required.

Finally, patients may not be contemporaneously identified as having SSI. Delayed or missed mild SSI diagnosis may present when the patient arrives in clinic for review (often after 30 days postoperatively, or if this is telephonic, it may be apparent through the history). Patients with more severe infection usually will have presented at the emergency department by this point, but if missed can impact on morbidity. Management will focus on ensuring no ongoing infection and alleviating further complications.

Index tests in this setting will likely be used as a comparative to face-to-face, gold standard review at planned review points. Potential implications of digital remote follow up are early and avoidance of missed diagnosis within the 30-day window for the CDC criteria.

### 2.4 Rationale

Widespread technological innovation and adoption have been exponential in the 21^st^ century. Sophistication of the mobile phone now allows for instantaneous communication all over the planet. Users can even transfer image and video data in real time. In 2019, 88% of individuals in the UK were estimated to own a smartphone(10). Naturally, the use of technology in healthcare has too progressed. Telemedicine is the remote diagnosis and treatment of patients by use of technology(11). Coined in the 1970s, this concept has broadened with the advent of the smartphone and mobile data. Mobile health (mHealth) is a contemporary classification whereby healthcare is supported by the use of mobile devices(12). The use of telemedicine has enabled patients in isolated centres access to specialist review through transfer of medical imaging, and teleconsultations are coming into practice(13). In surgery, the process of postoperative care is changing with the introduction of telemedicine.

In 2020, the COVID-19 pandemic caused a countrywide lockdown in the UK, and many other countries all over the world. Elective operations were cancelled, expanding already lengthy waiting lists. This also posed a challenge for outpatient follow up, as patients requiring review would be at risk of COVID-19 in attending the hospital, and departments adapted to comply with social distancing regulations, limiting the number of people allowed in outpatient spaces at any one time.

Remote follow-up has the potential to reduce unnecessary clinic visits providing benefits for both patients and healthcare providers. The rationale of this review, therefore, is to synthesise the current available evidence for using telemedicine to diagnose or exclude SSI in the context of post-operative follow up.

### 2.5 Objectives

#### 2.5.1 Research Question

Primary: Is digital remote follow-up accurate for the diagnosis of surgical site infection? Secondary:

- What methods are used to facilitate SSI diagnosis?
- What are the limitations of telemedicine in SSI diagnosis?

This systematic review aims to assess the diagnostic accuracy of using telemedicine to identify SSI post-operatively. It will also aim to identify which methods are currently in use for this and any limitations of telemedicine methods in SSI diagnosis.

#### 2.5.2 Objectives

Primary: To determine the diagnostic accuracy of digital remote follow-up for the diagnosis of surgical site infection

Secondary objectives

- To determine what methods of digital remote follow-up have been used
- Evaluate the accuracy of different digital remote follow-up methods
- To determine limitations of digital remote follow-up

## 3 Methods

### 3.1 Protocol Development

This protocol and review have been developed using the Cochrane handbook for reviews of diagnostic test accuracy and the Preferred Reporting Items for Systematic Reviews and Meta-Analyses (PRISMA) statements(14, 15). In addition, this protocol is reported in line with the PRISMA statement for review protocols (PRISMA-P) which is attached in appendix one(15).

### 3.2 Criteria for considering studies for this review

#### 3.2.1 Types of studies

There will be no restrictions on inclusion based upon prospective or retrospective study designs, however given the nature of the novel interventions, it is unlikely that retrospective studies will be available. Study designs which result in measures of test accuracy will be included, including randomised and observational studies. There will be no limitations on study sample sizes, or quality to thoroughly synthesise the available literature, but this will be accounted for in a quality of evidence assessment.

All study types will be included, and a sub group analysis performed for direct, fully paired and randomised studies.

Narrative and systematic review articles, letters and opinion pieces will be excluded from this review, however the reference lists of review articles will be hand searched for completeness.

#### 3.2.2 Participants

All patients over 18, who have undergone a procedure involving surgical incisions and followed up in a postoperative pathway will be eligible. Studies involving children under the age of 18 years and written in language other than English will be excluded from the review.

The study setting will vary depending on whether the index or reference test is being examined. The index tests can be performed anywhere as they are remote, the reference will be in a secondary care setting, likely clinics.

The index and reference tests will be applied on clinical assessment and on suspicion of surgical wound infection, no prior tests are required.

#### 3.2.3 Index and Comparative Tests

The index test to be reviewed is digital remote follow-up. Telemedicine is the remote diagnosis or treatment of patients using communications technology, which encompasses the term digital remote follow-up and will be used as a synonym in this review. Specifically, this entails the use of photographs or video, either deferred or in real time, telephone review and/or instant messaging. All index test methods will be evaluated through subgroup analyses.

The comparative tests are face to face review with the patient to directly observe the wound and obtain a history.

#### 3.2.4 Target Condition

Surgical site infection as defined by CDC is the target condition for this review; infection within 30 days of surgery or within 90 days if an implant is left in place. This is further discussed in section 1.1. Some studies may categorise this further into superficial, deep and organ/space SSI (more details in appendix two).

#### 3.2.5 Reference Standards

The same diagnostic criteria will be used for both tests. The gold standard for assessment of SSI is the US CDC criteria which clearly define indication of infection. Other possible methods are the ASEPSIS score with infection clearly define at a score of 21 or greater, and bluebelle WHQ. The latter has been suggested with an infection cut off score of 6-8. All methods of remote diagnosis will be extracted and a sub-group analysis performed for each type.

### 3.3 Search Methods for Identification of Studies

#### 3.3.1 Electronic searches

Medline, Embase, CENTRAL and CINAHL databases will be searched from inception to the current date. Additional resources will be identified through google scholar.

#### 3.3.2 Searching other sources

Additional searches will be conducted through handsearching the reference lists of included articles and excluded review articles. An example search strategy is provided in appendix five.

### 3.4 Data Collection and Analysis

#### 3.4.1 Selection of Studies

Search results will be deduplicated and uploaded to the specialised online review tool, Rayyan. Study titles and abstracts will be screened by two investigators, RL and MS, independently. Any disagreement will be resolved by consensus decision from a third investigator. Articles included at this stage will be retrieved for full text screening, again by two authors acting independently. Those included after full text screening will go on to data extraction.

#### 3.4.2 Data extraction and management

Data extraction will be into a bespoke designed spreadsheet (Microsoft Excel), hosted remotely and updated in real-time. Information on study design, country of origin, participant age and gender, sample size, surgery performed, drop out rates, time to follow-up and type of remote follow-up (photograph / video / telephone / other) will be extracted. Details on infection rates for remote and face to face methods as well as sensitivity and specificity of diagnosis will also be extracted. If information is available on specifics of superficial, deep and organ/space SSI rates, these data will also be extracted.

#### 3.4.3 Assessment of methodological quality

Methodological quality will be assessed using the QUADAS-2 tool (appendix four)(16). Due to the nature of varying study design, systematic reviews of diagnostic test accuracy are often prone to heterogenous results. In 2003, the quality assessment of diagnostic studies (QUADAS) tool was developed. This has since been revised as QUADAS-2 and is recommended for use in such reviews by the Agency for Healthcare Research and Quality, Cochrane Collaboration, and the UK National Institute for Health and Clinical Excellence(16).

Two authors will assess each manuscript as per the QADAS-2 tool, with a third independent author providing consensus review when discrepancies occur. Further details on the QUADAS-2 process can be found in appendix four.

#### 3.4.4 Statistical analysis and data synthesis

SSI diagnosed using any diagnostic criteria as part of a face-to-face review will be the reference standard. The patient will be the unit of analysis. Forest plots and receiver operating characteristic (ROC) curves will be produced as part of the initial, exploratory analysis and used to display estimates of sensitivity and specificity of the included studies.

Summary measures of sensitivity and specificity will be produced using a bivariate model for meta-analysis, if there are sufficient studies.

##### Receiver Operator Characteristic Curves

Summary receiver operator characteristic (sROC) curves will be plotted using each study included as a data point. Confidence regions will also be calculated. Plots will be produced using metaDTA(17).

##### Sensitivity Analysis

Sensitivity analysis based upon risk of bias will be evaluated. Studies with a high or unclear risk of bias identified by the QADAS-2 tool will be excluded in a separate analysis.

##### Assessment of Heterogeneity

Several sources of potential heterogenetiy have been identified, and their effects will be investigated through the use of subgroup analysis and metaregression. The diagnostic criteria used to diagnose SSI (CDC, ASEPSIS, Bluebelle WHQ or others) may influence the test accuracy as different tools have been found to have poor correlation(18).

## 4 Discussion

Integration of telemedicine has multiplied in recent years, exponentially so in response to the COVID-19 pandemic. One prospective application is remote diagnosis of SSI. Consolidation of telemedicine in surgical practice has the potential to reduce cost to both patient and care provider, as well as improved time implications for both parties. This would also reduce unnecessary visits to hospital clinic with a healthy surgical wound, an important consideration during a pandemic. To avoid further complications however and allow for confidence in diagnosis, telemedicine must be accurate in the detection of SSI. Previous studies have shown erythema detection to be difficult in review of wound images compare to face to face, which may influence a diagnosis of infection(19). This review aims to comprehensively examine the accuracy for all methods of remote diagnosis of SSI thereby enabling evidence-based decision making on remote reviews of post-operative patients.

## Data Availability

No data is produced in this protocol

## 6 Appendix / Supporting Information

### 6.1 PRISMA-P (Preferred Reporting Items for Systematic review and Meta-Analysis Protocols) 2015 Checklist: recommended items to address in a systematic review protocol(15) SSI Classification

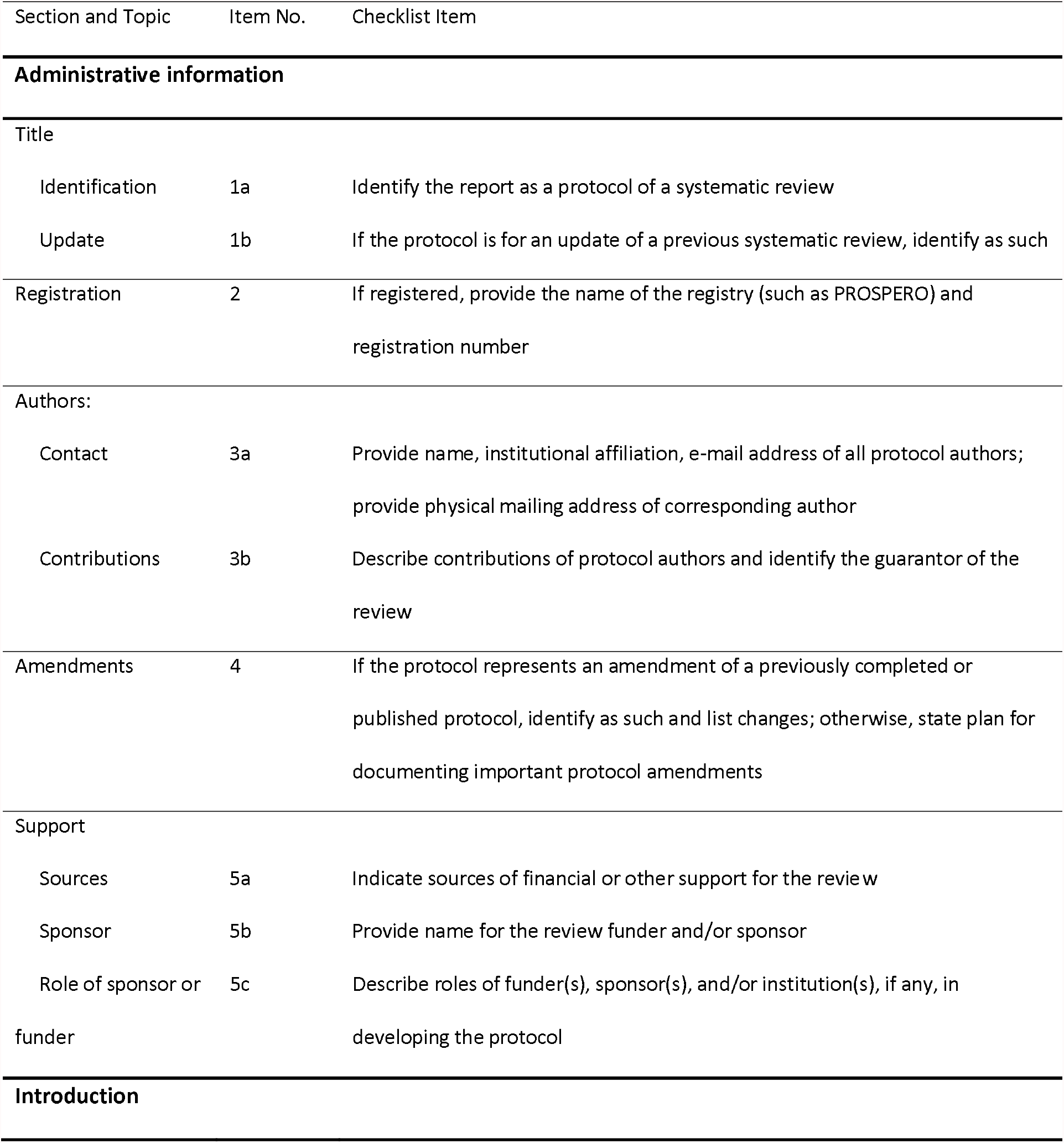
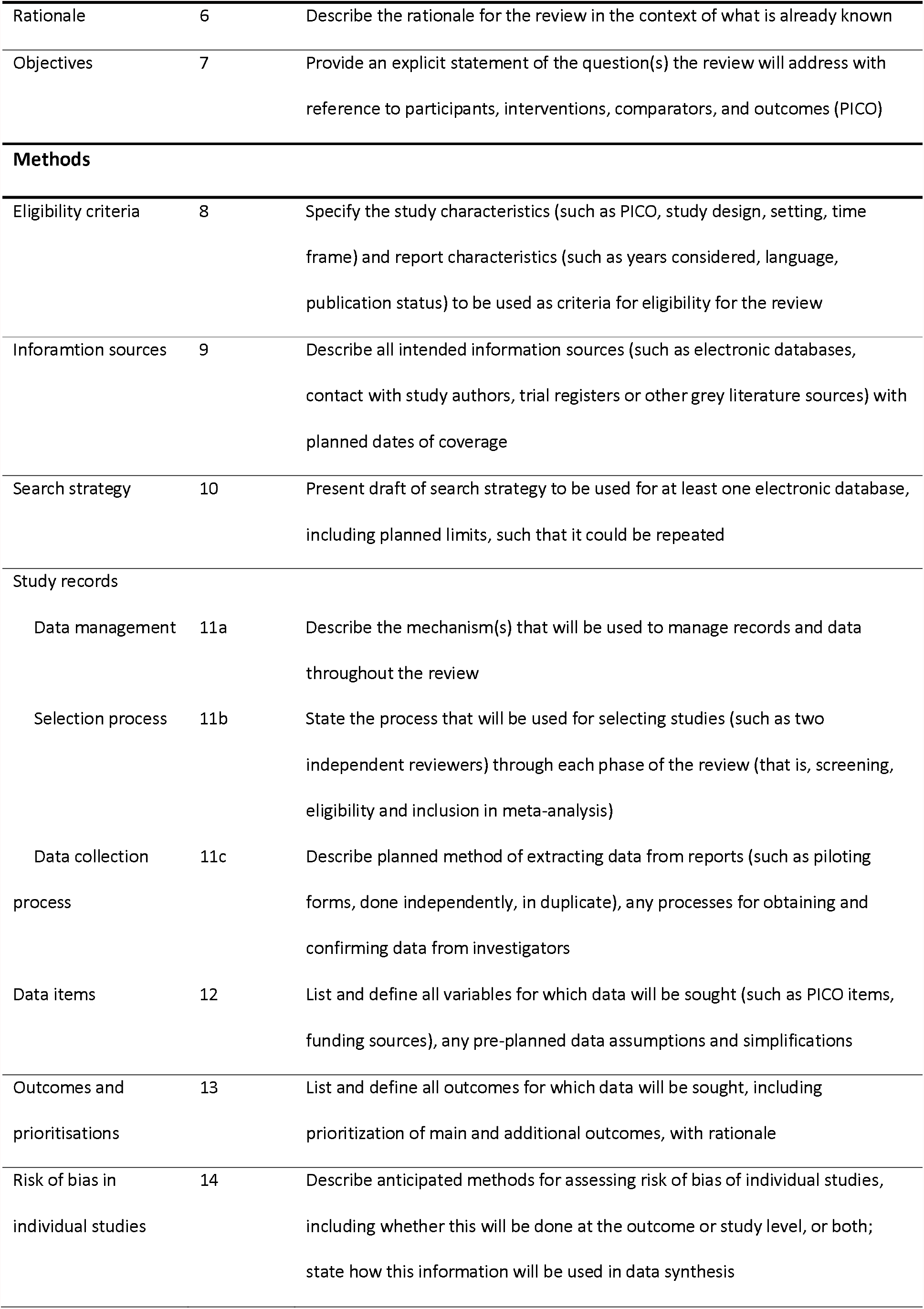
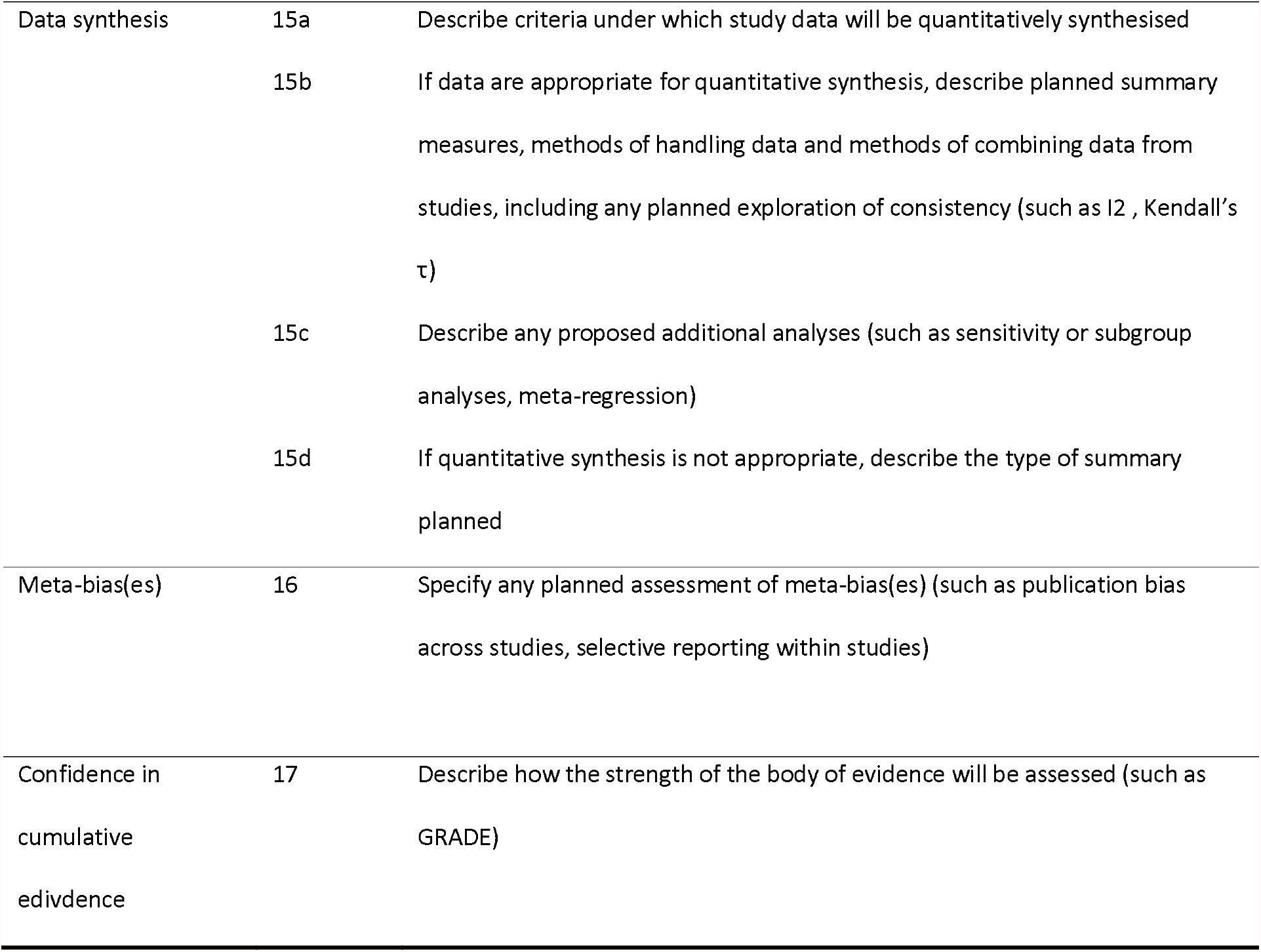

6.1 PRISMA-P (Preferred Reporting Items for Systematic review and Meta-Analysis Protocols) 2015 Checklist: recommended items to address in a systematic review protocol(15)
| Section and Topic | Item No. | Checklist Item |
| --- | --- | --- |
| **Administrative information** |  |  |
| Title |  |  |
| Identification | 1a | Identify the report as a protocol of a systematic review |
| Update | 1b | If the protocol is for an update of a previous systematic review, identify as such |
| Registration | 2 | If registered, provide the name of the registry (such as PROSPERO) and registration number |
| Authors: |  |  |
| Contact | 3a | Provide name, institutional affiliation, e-mail address of all protocol authors; provide physical mailing address of corresponding author |
| Contributions | 3b | Describe contributions of protocol authors and identify the guarantor of the review |
| Amendments | 4 | If the protocol represents an amendment of a previously completed or published protocol, identify as such and list changes; otherwise, state plan for documenting important protocol amendments |
| Support |  |  |
| Sources | 5a | Indicate sources of financial or other support for the review |
| Sponsor | 5b | Provide name for the review funder and/or sponsor |
| Role of sponsor or funder | 5c | Describe roles of funder(s), sponsor(s), and/or institution(s), if any, in developing the protocol |
| **Introduction** |  |  |
| Rationale | 6 | Describe the rationale for the review in the context of what is already known |
| Objectives | 7 | Provide an explicit statement of the question(s) the review will address with reference to participants, interventions, comparators, and outcomes (PICO) |
| **Methods** |  |  |
| Eligibility criteria | 8 | Specify the study characteristics (such as PICO, study design, setting, time frame) and report characteristics (such as years considered, language, publication status) to be used as criteria for eligibility for the review |
| Information sources | 9 | Describe all intended information sources (such as electronic databases, contact with study authors, trial registers or other grey literature sources) with planned dates of coverage |
| Search strategy | 10 | Present draft of search strategy to be used for at least one electronic database, including planned limits, such that it could be repeated |
| Study records |  |  |
| Data management | 11a | Describe the mechanism(s) that will be used to manage records and data throughout the review |
| Selection process | 11b | State the process that will be used for selecting studies (such as two independent reviewers) through each phase of the review (that is, screening, eligibility and inclusion in meta-analysis) |
| Data collection process | 11c | Describe planned method of extracting data from reports (such as piloting forms, done independently, in duplicate), any processes for obtaining and confirming data from investigators |
| Data items | 12 | List and define all variables for which data will be sought (such as PICO items, funding sources), any pre-planned data assumptions and simplifications |
| Outcomes and prioritisations | 13 | List and define all outcomes for which data will be sought, including prioritization of main and additional outcomes, with rationale |
| Risk of bias in individual studies | 14 | Describe anticipated methods for assessing risk of bias of individual studies, including whether this will be done at the outcome or study level, or both; state how this information will be used in data synthesis |
| Data synthesis | 15a | Describe criteria under which study data will be quantitatively synthesised |
| | 15b | If data are appropriate for quantitative synthesis, describe planned summary measures, methods of handling data and methods of combining data from studies, including any planned exploration of consistency (such as I<sup>2</sup>, Kendall's $\tau$) |
|  | 15c | Describe any proposed additional analyses (such as sensitivity or subgroup analyses, meta-regression) |
|  | 15d | If quantitative synthesis is not appropriate, describe the type of summary planned |
| Meta-bias(es) | 16 | Specify any planned assessment of meta-bias(es) (such as publication bias across studies, selective reporting within studies) |
| Confidence in cumulative evidence | 17 | Describe how the strength of the body of evidence will be assessed (such as GRADE) |

### 6.2 SSI Classification

#### Superficial incisional infection

Defined as a surgical site infection occurring within 30 days of surgery and involves only the skin or subcutaneous tissue of the incision, and meets at least one of the following criteria:

Criterion 1: Purulent drainage from the superficial incision.

Criterion 2: The superficial incision yields organisms from the culture of aseptically aspirated fluid or tissue, or from a swab and pus cells are present.

Criterion 3: At least two of the following symptoms and signs:

- pain or tenderness
- localised swelling
- redness
- heat

AND a. the superficial incision is deliberately opened by a surgeon to manage the infection, unless the incision is culture-negative

OR b. the clinician diagnoses a superficial incisional infection.

Note: Stitch abscesses are defined as minimal inflammation and discharge confined to the points of suture penetration, and localised infection around a stab wound. They are not classified as surgical site infections.

#### Deep incisional infection

Defined as a surgical site infection involving the deep tissues (i.e. fascial and muscle layers) that occurs within 30 days of surgery if no implant is in place, or within 90 days if an implant is in place and the infection appears to be related to the surgical procedure, and meets at least one of the following criteria:

Criterion 1: Purulent drainage from the deep incision but not from the organ/space component of the surgical site.

Criterion 2: The deep incision yields organisms from the culture of aseptically aspirated fluid or tissue, or from a swab and pus cells are present.

Criterion 3: A deep incision that spontaneously dehisces or is deliberately opened by a surgeon when the patient has at least one of the following symptoms or signs (unless the incision is culture negative):

- fever (>38°C)
- localised pain or tenderness

Criterion 4: An abscess or other evidence of infection involving the deep incision that is found by direct examination during re-operation, or by histo-pathological or radiological examination.

Criterion 5: Diagnosis of a deep incisional surgical site infection by an attending clinician.

Note: An infection involving both superficial and deep incision is classified as deep incisional SSI unless there are different organisms present at each site.

#### Organ/space infection

Defined as a surgical site infection involving any part of the anatomy (i.e. organ/space), other than the incision, opened or manipulated during the surgical procedure, that occurs within 30 days of surgery if no implant is in place, or within 90 days if an implant is in place and the infection appears to be related to the surgical procedure, and meets at least one of the following criteria:

Criterion 1: Purulent drainage from a drain that is placed through a stab wound into the organ/space.

Criterion 2: The organ/space yields organisms from the culture of aseptically aspirated fluid or tissue, or from a swab and pus cells are present.

Criterion 3: An abscess or other evidence of infection involving the organ/space that is found by direct examination, during re-operation, or by histo-pathological or radiological examination.

Criterion 4: Diagnosis of an organ/space infection by an attending clinician

Note: 1. Occasionally, an organ/space infection drains through the incision. Such infection generally does not require re-operation and is considered to be a complication of the incision, and is therefore classified as a deep incisional infection.

2. Where doubt exists, refer to the Definitions of specific site of organ/space infection to determine if the organ/space infection meets the definition

### 6.3 Asepsis Score

Scores awarded for ASEPSIS components
|  | Score |
| --- | --- |
| **Wound characteristic** |  |
| Serous exudates | 3 |
| Erythema | 3 |
| Purulent exudates | 6 |
| Separation of wound edges | 6 |
| **Additional treatment** |  |
| Postoperative Antibiotics | 10 |
| Abscess drainage | 5 |
| Wound debridement | 10 |
| Isolation of bacteria | 10 |
| Prolonged stay/admission to hospital | 5 |

**An ASEPSIS score of $\geq 21$ is taken as indicating the presence of infection, whilst a score of ≤**

### 6.4 QUADAS-2

The process occurs in four phases;

#### 1. Statement of the review question

This involves a summary of the review at hand in the form of patients, index tests, reference standards and target condition, details of which can be found in sections 2.2.2 – 2.2.5.

#### 2. Validation of QUADAS-2 tool

Both authors will utilise the tool in two example papers to be included as suggested in the QUADAS-2 supporting material. Agreement of 80% or more across all categories will be taken as sufficient and QUADAS-2 taken forward for assessment of all studies. Insufficient agreement will lead to further refinement of the tool through addition or omitting signalling questions and the process repeated until agreement is satisfactory.

#### 3. Flow diagram

The primary study’s flow diagram will be taken (or synthesised if not provided) to facilitate judgments of risk of bias in phase four. This will provide information on the method of patient recruitment, the order of test execution, the number of patients undergoing the index test and reference standard

#### 4. Judgments on bias and applicability

##### Risk of Bias

The tool itself comprises four separate domains, the first section of each concerns bias and has three sections.

- Information used to support the risk of bias judgment; to make the rating transparent and can aid discussion between independent review authors
- Signalling questions; presented to assist judgements. Answered ‘yes’, ‘no’, or ‘unclear’ so that ‘yes’ would indicate a low risk of bias.
- Judgement of the risk of bias; ‘low’, ‘high’, or ‘unclear’. If all signalling questions are answered ‘yes’ then risk of bias is to be judged as ‘low’. If any signalling question returns ‘no’ there would be potential for bias. The guidelines from phase two will assist authors in the judgement. ‘Unclear’ should only be used if insufficient data are reported to allow a judgement.

##### Applicability

These sections do not include signalling questions. Authors should record information on which the applicability judgement is made and then rate their concern that the study does not match the review question. This can be rated as ‘low’, ‘high’ or ‘unclear’. Again, ‘unclear’ is only to be used when insufficient data are available.

Guidance for completing the QUADAS-2 tool

#### Domain 1: Patient Selection

##### Risk of bias: Could the selection of patients have introduced bias?

- Was a consecutive or random sample of patients enrolled?
- Was a case control design avoided?
- Did the study avoid inappropriate exclusions?

Ideally studies should enrol all consecutive or a random sample of patients undergoing surgery, otherwise there is potential for bias. Inappropriate exclusions (difficult diagnosis) may skew estimates of diagnostic accuracy. Enrolment of patients with known SSI may also exaggerate diagnostic accuracy.

##### Applicability: Are there concerns that the included patients and setting do not match the review question?

Patients included in the study should match those in the review question to ensure applicability of results. This can be in terms of SSI severity, demographics, comorbidity, study setting.

#### Domain 2: Index Text

##### Risk of Bias: Could the conduct or interpretation of the index test have introduced bias?

- Were the index test results interpreted without knowledge of the results of the reference standard?
- If a threshold was used, was it pre-specified

The first signalling question refers to blinding, and the potential for bias with regards to testing order. If the index test is always conducted and interpreted prior to the reference standard, then this item can be rated ‘yes’.

The second question depends on the method of telemedicine used. If bluebelle WHQ is used in any of the methods this should be indicated what the cut off for SSI is.

##### Applicability: Are there concerns that the index test, it’s conduct, or interpretation differ from the review question?

Variations in test technology, execution or interpretation may affect estimates of it’s diagnostic accuracy. If these vary from those specified in the review question there may be concerns of applicability.

#### Domain 3: Reference Standard

##### Risk of Bias: Could the reference standard, its conduct, or its interpretation have introduced bias?

- Is the reference standard likely to correctly classify the target condition?
- Were the reference standard results interpreted without knowledge of the results of the index test?

What version of the reference standard was used? CDC / ASEPSIS / Bluebelle / other? How likely is that to have correctly identify SSI?

Potential for bias is related to the potential influence of prior knowledge on the interpretation of the reference standard, similar to that in domain two.

##### Applicability: Are there concerns that the target condition as defined by the reference standard does not match the question?

The target condition, SSI, defined by the reference standard, may differ from the SSI specified in the review question.

#### Domain 4: Flow and Timing

##### Risk of Bias: Could the patient flow have introduced bias?

- Was there an appropriate interval between index test and reference standard?

Ideally results on the index test and reference standard are collected on the same patient at the same time. Delay or initiation of treatment between one of the two tests may result in misclassification due to recovery or deterioration in SSI criteria.

- Did all patients receive the same reference standard?

Verification bias occurs when not all the study patients receive confirmation of diagnosis by the same reference standard.

- Were all patients included in the analysis?

All patients recruited into the study should be included in the analysis. If the number of patients enrolled differs from the number of patients included in the 2×2 table of results, there is potential for bias.

##### Presentation of QUADAS-2 results

Overall generalisable statements of ‘low’ or ‘high risk of bias’ will not be used unless a study is judged to be ‘low’ across all domains or if the study has been judged ‘high’ or ‘unclear’ in one or more domains it may be at risk of bias or as having concerns regarding applicability.

Results will be presented in the following cross tabulation whereby ‘+’ indicates high risk or concern, ‘-’ indicates low risk or concern and ‘?’ indicates unclear.

**Table 1:** Representation of tabulation for QUADAS-2 analysis on systematic review studies.

| Study | Risk of bias |  |  |  | Applicability Concerns |  |  |
| --- | --- | --- | --- | --- | --- | --- | --- |
|  | Patient Selection | Index Test | Reference Standard | Flow and Timing | Patient Selection | Index Test | Reference Standard |
| Study 1 | + / - / ? | + / - / ? | + / - / ? | + / - / ? | + / - / ? | + / - / ? | + / - / ? |
| Study 2 | + / - / ? | + / - / ? | + / - / ? | + / - / ? | + / - / ? | + / - / ? | + / - / ? |
| Study 3 | + / - / ? | + / - / ? | + / - / ? | + / - / ? | + / - / ? | + / - / ? | + / - / ? |
| ... | + / - / ? | + / - / ? | + / - / ? | + / - / ? | + / - / ? | + / - / ? | + / - / ? |

Recommended quality items derived from QUADAS tool

1. Was the spectrum of patients representative of the patients who will receive the test in practice? (representative spectrum)
2. Is the reference standard likely to classify the target condition correctly? (acceptable reference standard)
3. Is the time period between reference standard and index test short enough to be reasonably sure that the target condition did not change between the two tests? (acceptable delay between tests)
4. Did the whole sample or a random selection of the sample, receive verification using the intended reference standard? (partial verification avoided)
5. Did patients receive the same reference standard irrespective of the index result? (differential verification avoided)
6. Was the reference standard independent of the index test (i.e the index test did not form part of the reference standard) incorporation avoided)
7. Were the reference standard results interpreted without knowledge of the results of the index test? (index test results blinded)
8. Were the index test results interpreted without knowledge of the results of the reference standard (reference standard results blinded)
9. Were the same clinical data available when test results were interpreted as would be available when the test is used in practice? (relevant clinical information)
10. Were uninterpretable/intermediate test results reported? (uninterpretable results reported)
11. Were withdrawals from the study explained? (withdrawals explained) Additional items
12. Is the technology of the index test unchanged since the study was carried out?
13. Did the study provide a clear definition of what was considered to be a positive result?
14. Were data on observer variation reported and within an acceptable range?
15. Was treatment withheld until both the index test and reference standard were performed?
16. Were objectives pre-specified?

### 6.5 Example Search Strategy for Medline or Embase

(exp (Telemedicine)/ OR (remote consultation).mp OR (teleconsultation).mp OR (teleconsultation).mp OR (mobile health).mp OR (telehealth).mp OR (ehealth).mp OR (mhealth).mp OR (e*health).mp OR (e health).mp OR (m*health).mp OR (m health).mp OR (telephon*).mp OR (photograph*).mp OR (video*).mp OR (mobile app*).mp)

AND

(exp (surgical wound infection)/ OR (surgical wound dehiscence).mp OR surgical site infection).mp OR (postoperative infection).mp OR (SSI).mp OR (wound infection).mp OR (surgical wound complication).mp OR (post-surgical infection).mp OR (post operative infection).mp OR (post-operative infection).mp)

